# Multivariate GWAS of Alzheimer’s disease CSF biomarker profiles implies GRIN2D in synaptic functioning

**DOI:** 10.1101/2022.08.02.22278185

**Authors:** Alexander Neumann, Olena Ohlei, Fahri Küçükali, Isabelle J Bos, Stephanie Vos, Dmitry Prokopenko, Betty M Tijms, Ulf Andreasson, Kaj Blennow, Rik Vandenberghe, Philip Scheltens, Charlotte E Teunissen, Sebastiaan Engelborghs, Giovanni B Frisoni, Oliver Blin, Jill C Richardson, Régis Bordet, Alberto Lleó, Daniel Alcolea, Julius Popp, Christopher Clark, Gwendoline Peyratout, Pablo Martinez-Lage, Mikel Tainta, Richard JB Dobson, Cristina Legido-Quigley, Christine Van Broeckhoven, Rudolph E Tanzi, Mara ten Kate, Christina M Lill, Frederik Barkhof, Simon Lovestone, Johannes Streffer, Henrik Zetterberg, Pieter Jelle Visser, Kristel Sleegers, Lars Bertram, EMIF-AD & ADNI study group

## Abstract

Genome-wide association studies (GWAS) of Alzheimer’s disease (AD) have identified several risk loci, but many remain unknown. Cerebrospinal fluid (CSF) biomarkers may aid in gene discovery and we previously demonstrated that six CSF biomarkers (β-amyloid, total/phosphorylated tau, NfL, YKL-40, and neurogranin) cluster into five principal components (PC), each representing statistically independent biological processes. Here, we aimed to: 1. identify common genetic variants associated with these CSF profiles; 2. assess the role of associated variants in AD pathophysiology and 3. explore potential sex differences. We performed GWAS for each of the five biomarker PCs in two multi-center studies (EMIF-AD and ADNI). In total, 973 participants (n=205 controls, n=546 mild cognitive impairment, n=222 AD) were analyzed for 7,433,949 common SNPs and 19,511 protein-coding genes. Structural equation models tested whether biomarker PCs mediate genetic risk effects on AD, and stratified and interaction models probed sex-specific effects. Five loci showed genome-wide significant association with CSF profiles, two were novel (rs145791381 and *GRIN2D*) and three were previously described (*APOE*, *TMEM160B* and *CHI3L*). *GRIN2D* was associated with synaptic functioning, whereas rs145791381 was associated with biomarker evidence of inflammation. Mediation tests indicated that variants in *APOE* are associated with AD status via processes related to amyloid and tau pathology, while markers in *TMEM106B* and *CHI3L* are associated with AD only via neuronal injury/inflammation. Additionally, seven loci showed sex-specific associations with AD biomarkers. These results suggest that pathway and sex-specific analyses can improve our understanding of AD genetics and may contribute to precision medicine.

## Introduction

Alzheimer’s disease (AD) is a genetically complex disorder to which various pathophysiological processes are thought to contribute. Amyloid and tau pathology are the most well-known, but also other processes, such as inflammation and cholesterol metabolism, among many others, play important roles in disease development as well.^1^ Different risk factors may affect AD development by different mechanisms; therefore, patients may develop AD due to different combinations of causes and pathways. Accurately identifying and distinguishing which molecular mechanisms play the lead role on an individual basis is therefore crucial for etiological research, but also for clinical diagnosis, prognosis and future therapeutic approaches.

Cerebrospinal fluid (CSF) biomarkers can provide insights into disease mechanisms, often before symptoms fully develop.^2^ We have previously demonstrated the utility of linearly combining different AD CSF biomarkers into five statistically independent components, which likely represent different disease processes and which may be more informative than analyzing each CSF trait separately.^3^ Specifically, we had applied principal component analysis (PCA) to data for six CSF biomarkers collected in two cohorts: the European Medical Information Framework for Alzheimer’s Disease Multimodal Biomarker Discovery (EMIF-AD MBD) study^4^ and the Alzheimer’s Disease Neuroimaging Initiative (ADNI)^5^.

The five principal components (PC) can be summarized as follows:^3^ the first PC loaded strongly on tau and phosphorylated tau (pTau), and moderately on neurogranin (Ng) and YKL-40. Tau is a marker of neurodegeneration, with pTau being a component of neurofibrillary tangles,^2,6^ Ng is a marker of synaptic functioning^7^, while YKL-40 is associated with neuronal inflammation and astroglial reaction.^2,8–10^ Thus, this PC likely represents tau pathology and associated degenerative processes, such as deficits in synaptic functioning and elevated inflammation (henceforth referred to as “tau pathology/degeneration” (PC1)). The second PC loads specifically on Aβ42 only (“Aβ Pathology” PC2), a very early and important marker of amyloid deposition in the brain.^2^ The third component loads strongly on neurofilament light chain (NfL), but also moderately on YKL-40, and can be interpreted as representing neuronal injury and the accompanying inflammatory response (“injury/inflammation” PC3), as NfL is a component of axons and its presence in CSF is a non-specific marker of neuronal damage.^6^ The fourth component loads on YKL-40 and only weakly on tau and NfL, and therefore can be regarded as representing neuronal inflammation and astroglial reaction, not related to AD symptoms (“non-AD inflammation” PC4). Similarly, the fifth component loads strongly on Ng and weakly on tau, representing synaptic functioning mostly independent of the other biomarkers and AD symptoms (“non-AD synaptic functioning” PC5).

After establishing the component structure, we applied these to search for rare-variant associations using whole-exome sequencing (WES) in our previous study.^3^ This work led to the identification of six genes, in which rare variants were associated with the CSF PCs. Specifically, we identified associations between the injury/inflammation component (PC3) and rare variants in *IFFO1*, *DTNB*, *NLRC3* and *SLC22A10*, as well as between the non-AD synaptic functioning component (PC5) and rare variants in *GABBR2* and *CASZ*.^3^ Interestingly, rare-variant associations with AD risk were simultaneously reported for the *DTNB* locus in an independent project utilizing whole-genome sequencing in AD families and case-control datasets.^11^

In this study, we aimed to extend the previous analyses to investigate the role of common variants on the PCA-defined CSF biomarker profiles. While previous GWAS in the field have screened for common-variant associations with single biomarkers,^9,12–15^ to our knowledge no GWAS combining these CSF biomarkers in a multivariate framework has been performed to date. Multivariate analyses have the advantages of i) allowing a more robust (compared to univariate analyses) quantification of different disease pathways, resulting in increased statistical power^16,17^ and, ii) enabling to differentiate various possible mechanisms of action more precisely.

Secondary aims of our study include the identification of sex-specific effects and AD mediation pathways. AD is more prevalent in women, and CSF biomarkers differentially predict brain and cognitive changes depending on sex.^18,19^ Furthermore, genetic effects on CSF biomarkers may depend on sex as well, e.g. rs34331204 on chromosome 7p21 was found to have a male-specific association with neurofibrillary tangles.^20^ It is therefore prudent to investigate whether the component structure differs between sexes and whether associations of PCs with AD or with genetic predictors is sex-dependent. Finally, mediation analyses allow to gauge whether potential SNP effects on CSF biomarker profiles also affect AD risk.

## Methods

### Participants

The presented work is part of the EMIF-AD project, a consortium of European studies investigating the etiology of AD and AD biomarkers with the aim to improve prognosis and diagnosis.^4^ Participants included elderly individuals with cognitively unimpaired individuals, mild cognitive impairment (MCI) and AD type dementia. Both deep phenotyping (such as brain imaging and determination of CSF biomarkers) and genotyping (SNP arrays and WES) were performed on a large part of EMIF-AD participants.^21–23^ The current study utilizes the existing CSF biomarker and SNP array data and combines them with a range of statistical methods not previously employed on these data. Written informed consent was obtained for all assessment before the start of the study.^4^ The study was conducted in accordance to the Declaration of Helsinki and ethical approval was obtained from the Ethical Committee of the University of Lübeck, as well as local committees of consortium members.^4^ More details on the recruitment and phenotype ascertainment protocols used in the EMIF-AD dataset can be found in Bos et al.^4^

To increase the generalizability of effect estimates and to increase power to detect new associations, we performed all analyses jointly with equivalent CSF biomarker and SNP genotype data from the Alzheimer’s Disease Neuroimaging Initiative (ADNI).^5^ Data used in the preparation of this article were obtained from adni.loni.usc.edu. ADNI was launched in 2003 as a public-private partnership, led by Principal Investigator Michael W. Weiner, MD. The primary goal of ADNI has been to test whether serial magnetic resonance imaging (MRI), positron emission tomography (PET), other biological markers, and clinical and neuropsychological assessment can be combined to measure the progression of MCI and early AD.

The current study utilized two participant selection paradigms for analysis: first, we selected participants for whom observations for at least 4 out of the 5 biomarkers were available. In total, this yielded 1158 participants to construct and examine the biomarker PC scores (Table S1). Second, we only included participants with available SNP array data, who were unrelated and of European ancestry. This reduced the sample size to 973 participants (Table S2, see also Hong et al. for detailed selection methods^13^). Overall, both EMIF-AD and ADNI were comparable datasets of elderly participants, with a mean age at ascertainment of 69 and 75 years, respectively (Table S1). The distributions of diagnostic status were similar in both datasets as well, with approximately half of the sample diagnosed with MCI, while 25% presented either no cognitive impairment or with a diagnosis of AD (Table S1).

### Measures

#### Genotyping, imputation and quality control (QC)

SNP genotypes were determined using the Infinium Global Screening Array (GSA; Illumina, Inc., USA) at the Institute of Clinical Molecular Biology (UKSH, Campus-Kiel) in EMIF-AD and using Illumina’s Omni 2.5M or Human610-Quad arrays in ADNI. Autosomal SNPs in both GWAS datasets were processed with the same computational workflow^13^, including the imputation of untyped variants with MiniMac 3 using the HRC 1.1 reference panel^24^. Here, we only analyzed common SNPs (MAF>=0.01 per study) with sufficient imputation quality (R^2^>0.30) and SNPs within HWE (p < 5*10^−6^). Please see Hong et al. for a detailed description of the GWAS methods, QC criteria and processing pipeline.^13^ For X-chromosome specific methods, see supplementary methods.

#### CSF biomarkers and dementia symptoms

Biomarkers were derived from CSF, as obtained via lumbar puncture.^3,5,25^ For EMIF the V-PLEX Plus AbPeptidePanel 1 Kit was used to measure Aβ and in the case of tau, the INNOTEST ELISA was applied.^25^ In ADNI the Elecsys CSF immunoassay and a cobas e 601 analyzer assessed Aβ and tau concentration.^26^ For both cohorts ELISA was applied to assess NfL levels.^25,27^ Ng concentration was measured by ELISA in EMIF^25^ and by electrochemiluminescence in ADNI^28^. ELISA was used to measure YKL-40 levels in EMIF^25^ and LC/MRM-MS proteomics were applied in ADNI^29^. For the YKL-40 proteomics data we z-score standardized two ion frequencies with two peptide sequences each and averaged the values.

### Statistical analysis

#### CSF biomarker PCA, sex differences and AD associations

We computed five PCs across all participants with sufficient biomarker information. PCs were defined as described previously^3^ and assigned to specific functional domains, as described in the introduction. Briefly, we first applied a rank based inverse normal transformation (INT) within both studies to decrease extreme skewness of the observed biomarker levels and to z-score standardize the scale across studies.^30^ We then applied a PCA-based imputation approach using the missMDA package to impute missing values, so that excessive sample size losses and potential participation biases could be avoided.^31^ Using leave-one-out cross-validation, a similar five component structure could be consistently identified and derived in both studies. PCA scores were computed with the psych package.^32^ All analyses were performed in R 4.0.3^33^

We have not previously explored to which degree the component structure differs between sexes, or whether the resulting PCs show sex-dependent associations with dementia symptoms. We first repeated PCA in both males and females and compared loadings. We then tested whether mean PC levels differed between sexes. This was achieved by regressing PC scores on sex, adjusted for age, five genetic ancestry components, diagnostic status (dummy coding MCI and AD), and study (ADNI vs EMIF-AD).

In a last step, we used the PC scores as predictor of latent AD. Here, latent AD is defined as an underlying continuous normally distributed variable, representing a range of probability to either have no cognitive impairment, MCI or AD. Latent AD was estimated by item factor analysis.^34^ Accordingly, participants with low scores (below −1.47SD) are unlikely to display cognitive impairment, above −1.47SD and below 0.40SD are most likely to suffer from MCI and above 0.40SD have a high probability to be affected by AD. To account for potential sex differences, we also added a product term between PC scores and sex, coded as male = 0 and female = 1. The biomarker PC term can therefore be interpreted as the association of biomarker PC scores on latent AD in males and the interaction term as the female-specific effect, i.e., the difference between sexes. These analyses were adjusted for the same covariates as in the main analyses, i.e. age, genetic ancestry, and study. We applied a structural equation model (SEM) with a weighted least square mean and variance adjusted (WLSMV) estimator using Lavaan 0.6-9^35^ to estimate latent AD and regress it onto biomarker PCs, sex, their interaction and covariates.

##### GWAS and meta-analyses

We performed four sets of GWAS: main GWAS analysis (both sexes), male-only GWAS, female-only GWAS, and sex-interaction analyses. Within each analysis group, we performed five GWAS, one for each biomarker PC, separately for EMIF-AD and ADNI. For all GWAS, separate linear regression models were run in PLINK^36^ to all autosomal and X-linked SNPs passing QC. For the X-chromosomal analyses genotype, dosage for hemizygous males was coded as 2, to reflect the same dosage as homozygous females.^37^ The biomarker PC scores were treated as outcome, imputed SNP dosage (0-2 numbers of effect allele) as main predictor. In addition, we included sex and five PCs reflecting genetic ancestry as additional covariates in the regression models. Analyses of the ADNI dataset were additionally corrected for genotyping array. Lastly, GWAS results for the EMIF-AD and ADNI were meta-analyzed using the inverse variance weighting model implemented in METAL.^38^

In secondary analyses, we aimed to discover SNP effects exclusively found in one sex by running sex-specific GWAS. In the final model, we added a sex-interaction term, representing the difference between the SNP effect in females vs males. In addition to single variant analyses, we also estimated the aggregate effect of all SNPs within a protein-coding gene. These analyses were performed with MAGMA 1.08 ^39^ on the FUMA 1.3.7 platform with default settings.^40^ FUMA was also used to select independent genome-wide significant (p<5*10^−8^) SNPs for further mediation analyses.

#### Mediation analyses

Independent SNPs, which showed genome-wide significant association in any of the GWAS were further tested for mediation effects. Specifically, we examined whether these SNPs would affect latent AD via their influence on biomarker levels. To test this hypothesis, we applied a SEM to each SNP. In this SEM the genetic variant predicts all biomarker PCs, as well as latent AD directly. The biomarker PCs in turn also predict latent AD. See Figure 1 for a path diagram. Sex, age, five genetic ancestry components and study were predictors of both CSF biomarkers and latent AD, thus all mediation and direct pathways were statistically adjusted for these potential confounders. Paths from SNPs to a biomarker were multiplied with the path from biomarker to latent AD to obtain the mediation effect for that particular PC. We also summed all mediation pathways to obtain a total mediation estimate elicited by any biomarker. This biomarker mediation estimate was further summed with the direct effect to estimate the total effect. To aid the interpretation of the mediation magnitude, we also provide an estimate of the proportion of the mediated effect (i.e., biomarker mediation/total effect). However, this was only possible, when mediated and total effects pointed in the same direction. Additional context to the total effect is afforded by providing the variants’ effects on AD based on a large and independent previous case-control GWAS.^41^

**Figure 1:**
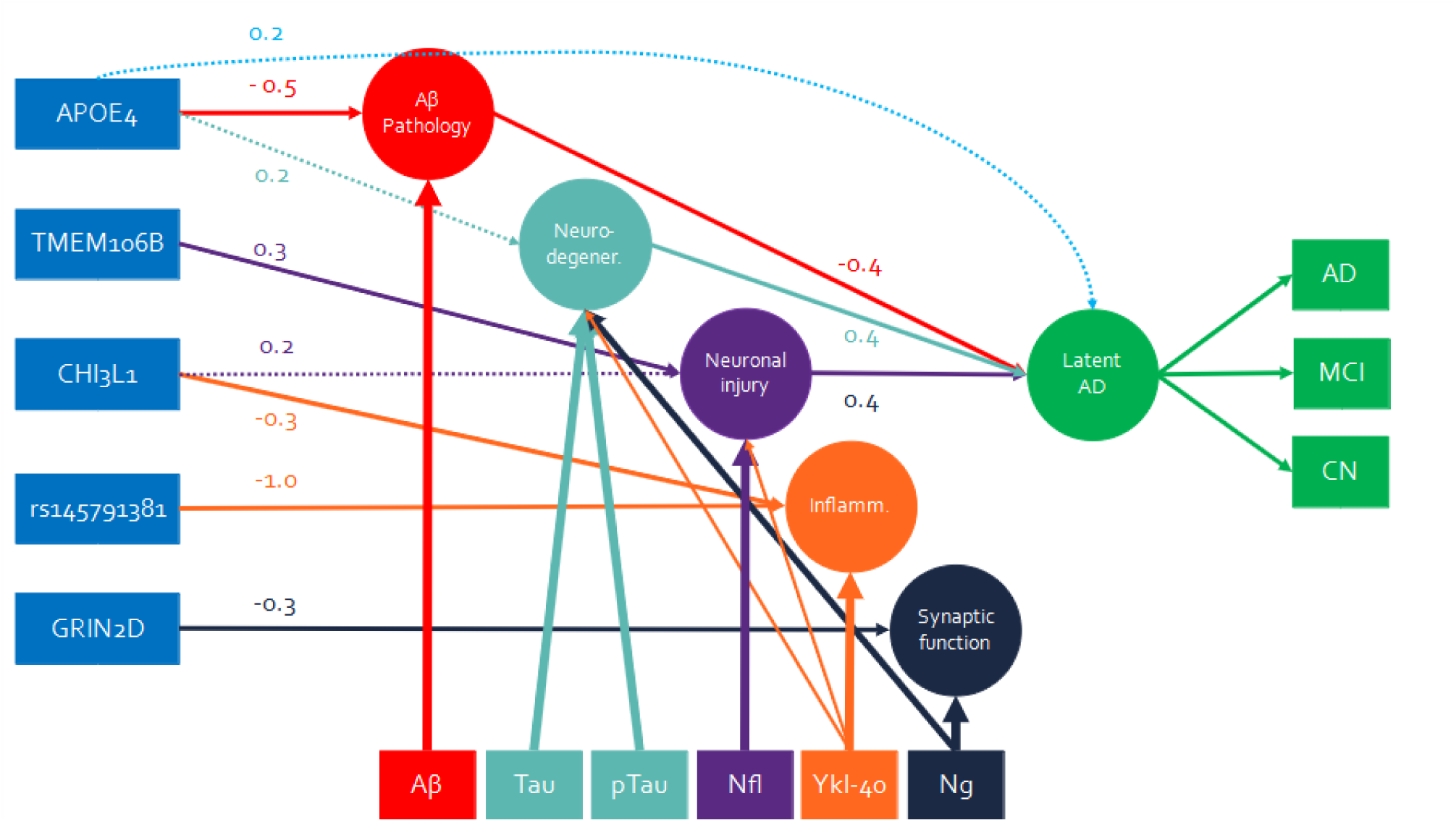
Path model of main findings. This path model summarizes the main effects mediation model. Circles indicate principal components or latent variables, rectangles represent observed variables. Arrows either indicate PC loadings or structural regression paths. Thicker lines correspond to stronger loadings, solid structural paths are genome-wide significant (p<5*10*^−8^) and dashed lines are suggestive (p<0.008). Coefficients indicate either the effect of one effect allele on a biomarker PC in SD, or the effect of one SD higher biomarker PC score on latent AD in SD. Note: all paths are adjusted for assessment age, sex, genetic ancestry and study but are omitted from figure.

**Figure 2:**
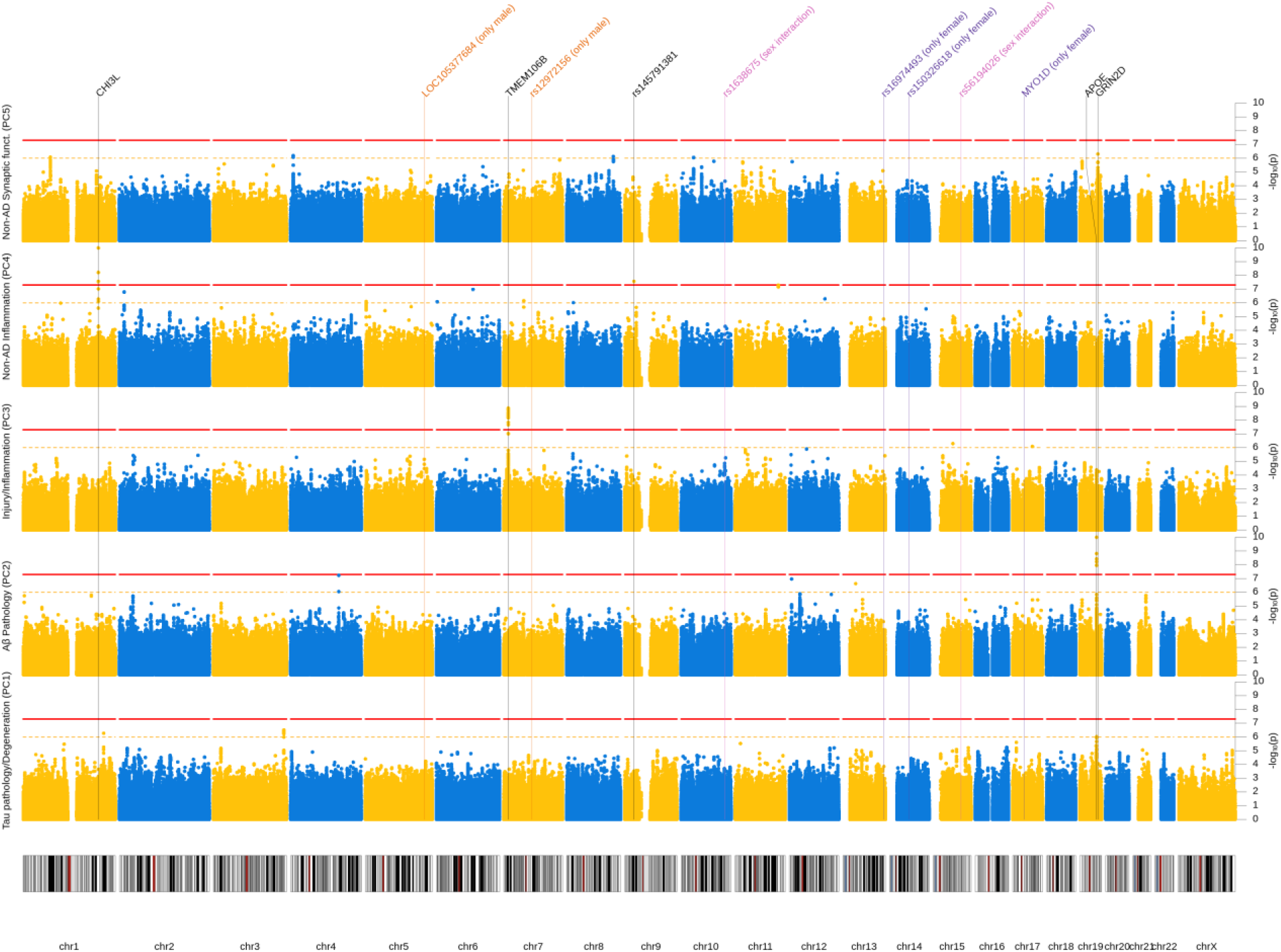
Manhattan plot (main effect model). Results from GWAS of five CSF biomarker PC across both sexes. Each row represents a different PC as outcome. X-axis represents each SNP and the y-axis the p-value of the SNP association with the outcome on a −log_10_ scale. All analyses were adjusted for sex, genetic ancestry and SNP array. Red line indicates genome-wide significance threshold (p=5*10^−8^). Yellow line indicates suggestive threshold (p=1*10^−6^). Vertical lines point towards genome-wide significant loci based on any model. P-values below 1*10^−10^ were winsorized to 1*10^−10^.

As some SNPs showed sex-dependent effects, we also ran a moderated mediation model to account for sex-specific mediation. This was achieved by adding a product term between SNP dosage and dummy variable for sex (female = 1, male = 0), and adding this interaction term as predictor of biomarker PCs and latent AD. If no mediation pathway differed nominally (p≥0.05) between sexes, main mediation model results are presented. Otherwise, male and female specific mediation estimates are provided, as estimated by the moderated mediation model. All mediation analyses were estimated with WLSMV in lavaan.^35^

#### Comparison to rare variant results

As outlined in the introduction, we previously identified several rare-variant associations using the same biomarker PCA approach in a subset of the EMIF-AD MBD individuals analyzed here. Specifically, this pertains to associations between *IFFO1*, *DTNB*, *NLRC3* and *SLC22A10* and the injury/inflammation component (PC3), as well as between *GABBR2* and *CASZ* and the non-AD synaptic functioning component (PC5).^3^ Here we examined, whether common variants – in addition to the rare-variants already identified – in these genes also show associations with the CSF biomarker PCs, using both single variant and gene-based tests as outlined above.

#### Multiple testing adjustment

To strike a balance between reliable inference and power, we present our findings as primary, secondary and tertiary results. The primary analyses in this study were the GWAS in the full dataset independent of sex. For SNP-based tests we apply the conventional genome-wide association threshold of p<5*10^−8^ and for gene-based tests we used Bonferroni’s method to adjust for 19,511 genes resulting in a threshold of p<2.3*10^−6^, as recommended by FUMA. The sex-specific analyses present additional tests of related (and non-independent) hypotheses, and, thus should be regarded as secondary and more exploratory analyses. For the mediation analyses we applied an alpha of 0.05/6=p<0.0083, adjusting for six potential mediation or direct pathways. See supplementary methods for full description of the multiple testing adjustment strategy.

## Results

### CSF biomarker PCs

Analogous to our previous study,^3^ the six AD CSF biomarkers tested here could be combined into five consistent components across datasets and analytical subsamples. In this study we extended our analyses to examine whether the CSF biomarker PCs’ loadings, their mean levels, or associations with latent AD differ by sex. Generally, PC loadings were consistent across males and females (Table 1). NfL loaded 0.06 higher on tau pathology/degeneration in females when compared to males (0.25 vs 0.19). A similar pattern was observed for pTau, which loaded 0.07 higher on Injury/Inflammation (0.18 vs 0.11) in women vs men. All other loading differences were below 0.04 and therefore classified as “indifferent” between sexes. Based on these observations we used the common loadings as estimated across both sexes for further analyses.

**Table 1:** PCA loadings. Component loadings of each biomarker (first column) on five principal components (column groups two to six) are displayed.

| Sample | Tau pathology/<br>Degeneration (PC1) | | | Aβ Pathology (PC2) | | | Injury/<br>Inflammation (PC3) | | | Non-AD<br>Inflammation (PC4) | | | Non-AD<br>Synaptic functioning (PC5) | | |
| --- | --- | --- | --- | --- | --- | --- | --- | --- | --- | --- | --- | --- | --- | --- | --- |
|  | Male<br>(n=601) | Female<br>(n=557) | All<br>(n=1158) | Male<br>(n=601) | Female<br>(n=557) | All<br>(n=1158) | Male<br>(n=601) | Female<br>(n=557) | All<br>(n=1158) | Male<br>(n=601) | Female<br>(n=557) | All<br>(n=1158) | Male<br>(n=601) | Female<br>(n=557) | All<br>(n=1158) |
| Tau | 0.86 | 0.87 | 0.87 | -0.05 | -0.07 | -0.06 | 0.23 | 0.25 | 0.23 | 0.26 | 0.24 | 0.24 | 0.27 | 0.27 | 0.27 |
| pTau | 0.91 | 0.89 | 0.91 | -0.08 | -0.07 | -0.08 | 0.11 | 0.18 | 0.14 | 0.19 | 0.23 | 0.20 | 0.26 | 0.28 | 0.26 |
| Aβ | -0.07 | -0.07 | -0.07 | 1.00 | 1.00 | 1.00 | -0.03 | -0.02 | -0.03 | -0.01 | 0.00 | 0.00 | 0.04 | 0.04 | 0.04 |
| NfL | 0.19 | 0.25 | 0.21 | -0.04 | -0.03 | -0.03 | 0.95 | 0.93 | 0.94 | 0.25 | 0.27 | 0.25 | 0.08 | 0.10 | 0.08 |
| YKL-40 | 0.31 | 0.33 | 0.32 | -0.01 | 0.01 | 0.00 | 0.30 | 0.33 | 0.32 | 0.88 | 0.87 | 0.88 | 0.17 | 0.17 | 0.17 |
| Ng | 0.49 | 0.51 | 0.51 | 0.06 | 0.07 | 0.07 | 0.11 | 0.12 | 0.10 | 0.18 | 0.19 | 0.18 | 0.84 | 0.83 | 0.83 |

While, the component structure was very similar between sexes in general, we observed differences in mean levels. When adjusting for age, diagnostic status and study, females showed 0.21SD (SE=0.06, p=6.3*10^−4^) higher scores on non-AD synaptic functioning. In contrast, injury/inflammation was −0.40SD (SE=0.40, p=2.7*10-^13^) lower in females. Tau pathology/degeneration, Aβ pathology and non-AD inflammation did not show differences in mean levels across sexes when accounting for multiple testing (p<0.05/5=0.01).

Finally, we examined the association of the biomarker PCs with latent AD, including potential sex interactions (Table S3). 1SD higher levels in tau pathology/degeneration or injury/inflammation were associated with 0.41SD and 0.40SD higher probability of MCI or AD (latent AD factor) in males. Females had a stronger association with 0.43SD and 0.44SD, respectively, but the difference was not significant (p>0.5). Higher brain Aβ accumulation is reflected in lower CSF Aβ values, therefore, higher Aβ pathology scores were associated with lower AD occurrence in both males and females (β=-0.34SD and β=-0.45), although the difference between sexes was not significant (p=0.13). Non-AD inflammation and non-AD synaptic functioning did not significantly associate with latent AD, when adjusting for five tests (i.e., all p>0.01). Due to lack of evidence for sex-differential associations all subsequent analyses are performed under the assumption that associations between PCs and latent AD are invariant across sexes.

### GWAS

In our GWAS analyses we tested 7,433,949 autosomal and X-linked SNPs. For gene-based tests, we assessed 19,511 protein-coding genes. For all outcomes and analyses, lambda was below 1.05 and QQ-plots showed no evidence of noteworthy genome-wide inflation (Figure S1, S1). GWAS results are visualized as Manhattan plots for main (Figure 3), sex stratified (Figure 4), sex interaction analyses (Figure S2) and gene-based tests (Figure S3). Results of independent SNPs showing genome-wide significance are summarized in Table 2. Their effect on AD risk by mediation analysis using CSF biomarker PCs are displayed in Table 3. Finally, gene-based results are depicted in Table 4.

**Figure 3:**
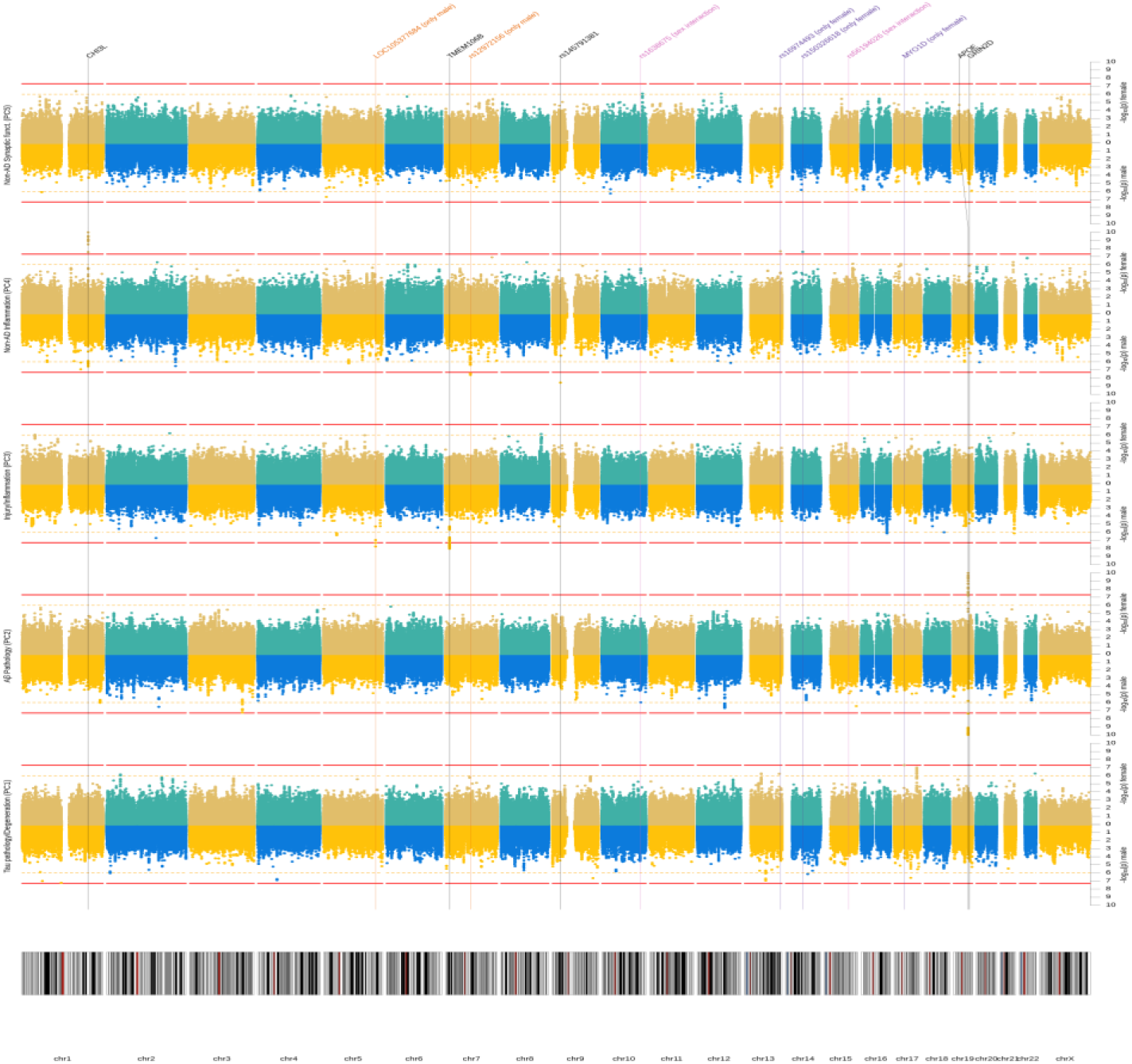
Manhattan plot (sex stratified). Results from GWAS of five CSF biomarker PC for males and females separately. Each row represents a different PC as outcome. Per outcome, results for males are depicted at the bottom and for females at the top. X-axis represents each SNP and the y-axis the p-value of the SNP association with the outcome on a −log_10_ scale. All analyses were adjusted for genetic ancestry and SNP array. Red line indicates genome-wide significance threshold (p=5*10^−8^). Yellow line indicates suggestive threshold (p=1*10^−6^). Vertical lines point towards genome-wide significant loci based on any model. P-values below 1*10^−10^ were winsorized to 1*10^−10^.

**Figure 4:**
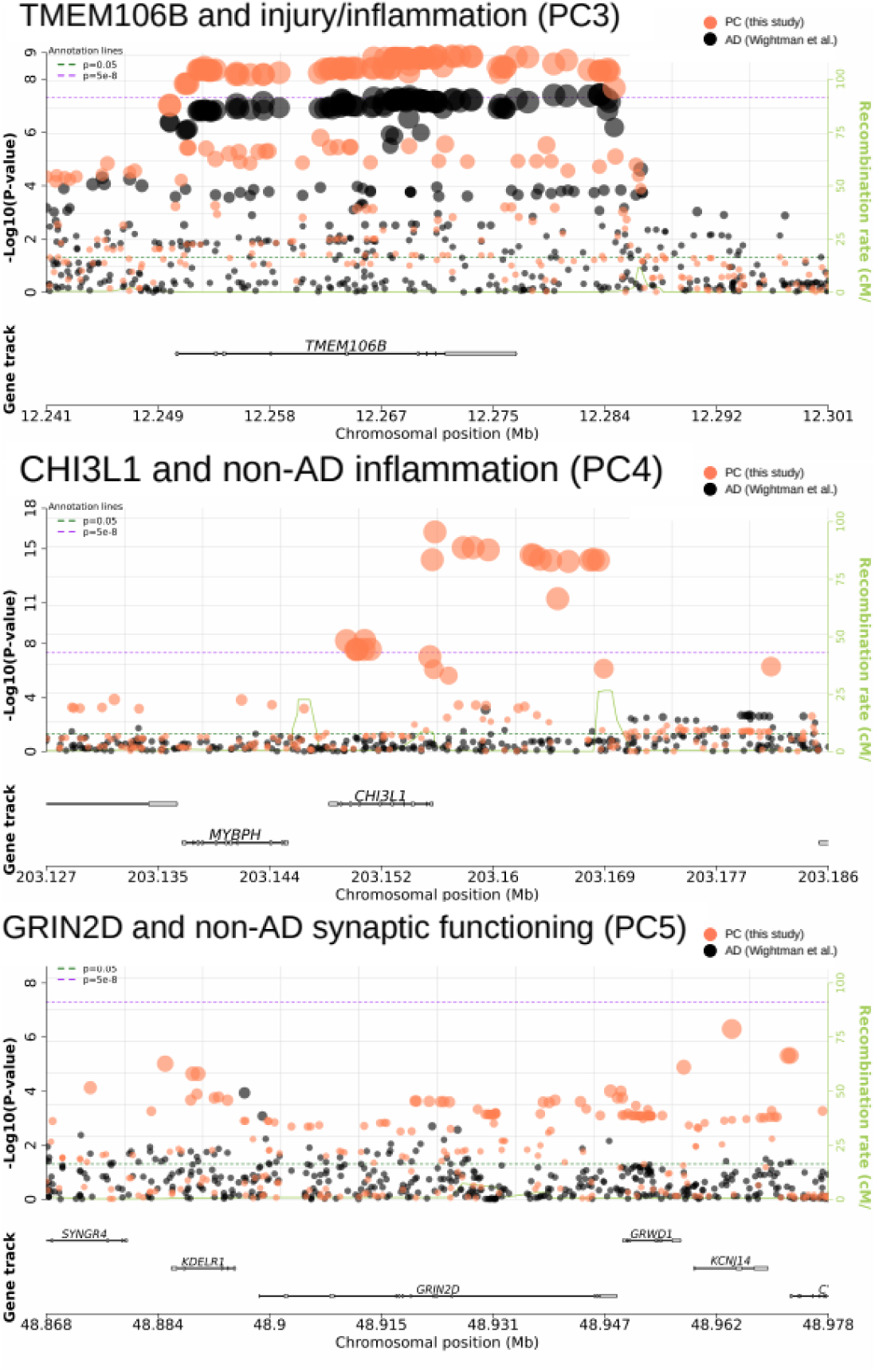
Regional plots for *TMEM106B*, *CHI3L1* and *GRIN2D*. Each plot displays the p-values of SNP associations in either *TMEM106B*, *CHI3L1* or *GRIN2D* loci. Statistics are derived from two studies. Orange dots represent p-values of association with biomarker PCs estimated in this study and back dots represent p-values of association with AD, as estimated in a separate GWAS on 1 million participants (Wightman et al., 2022). Regional plots were created with snpxplorer.net.

**Table 2:**
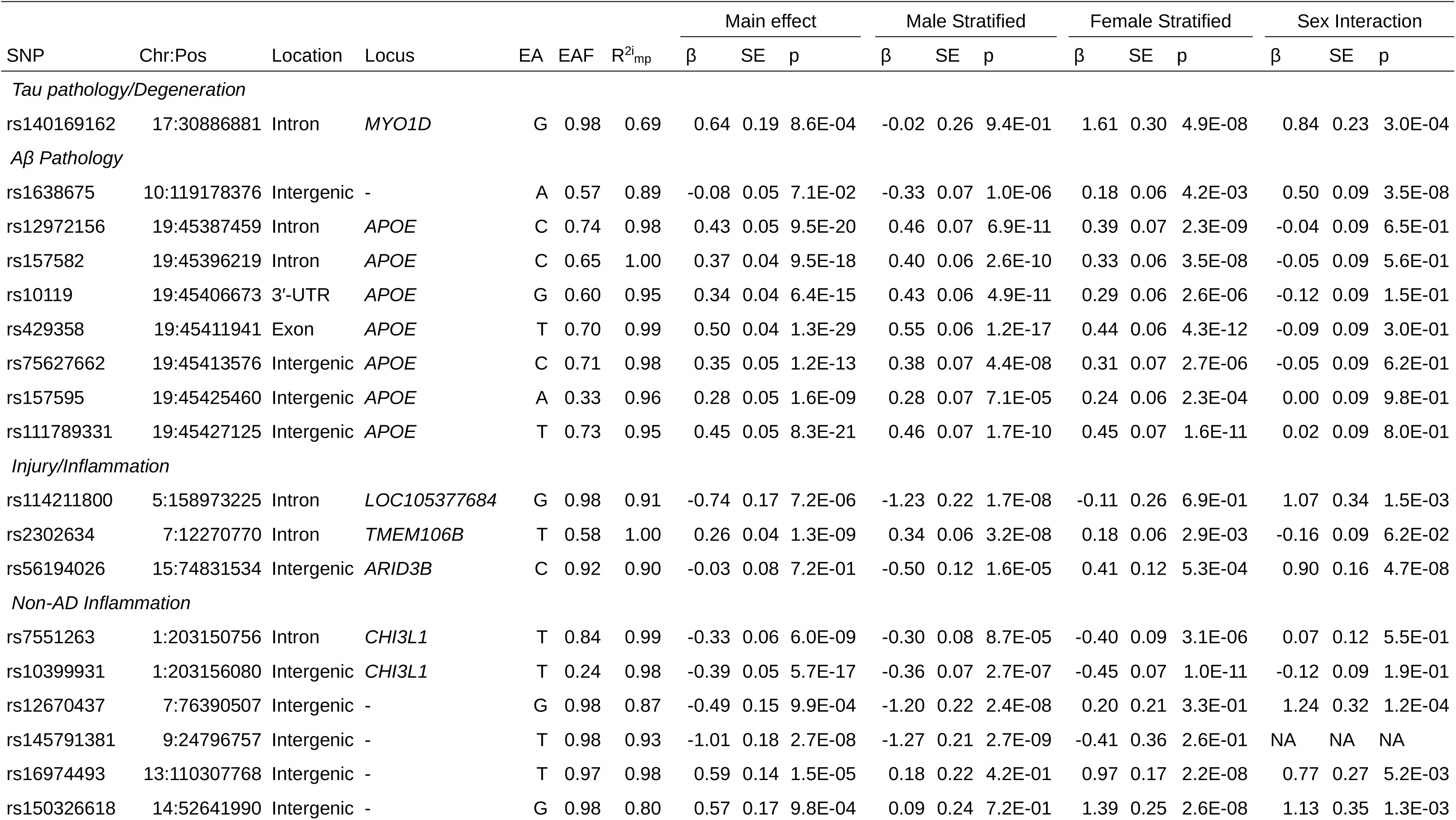
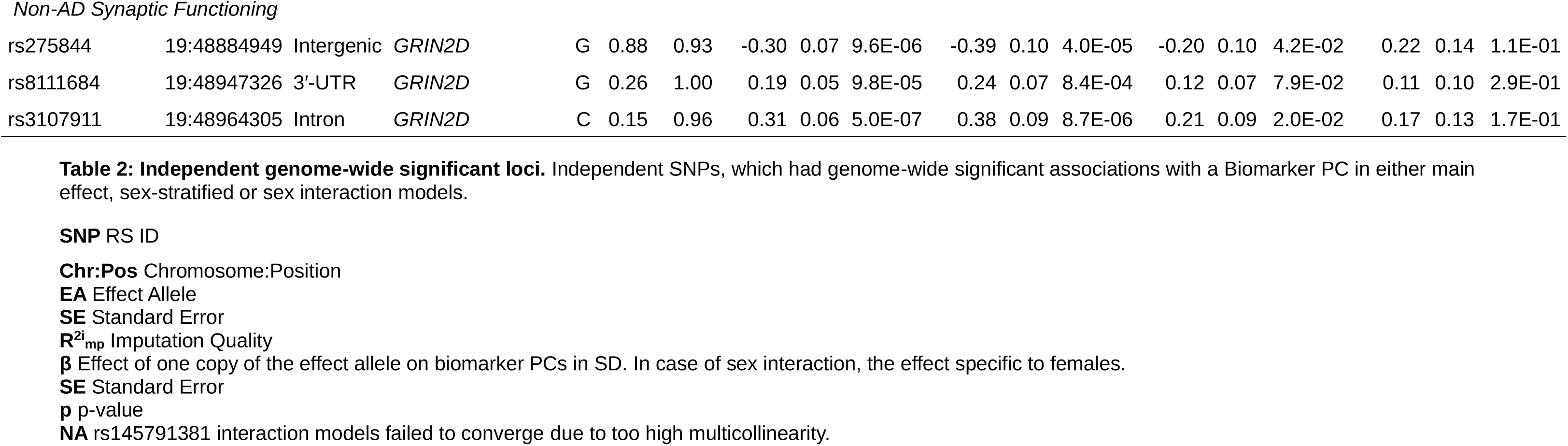
Independent genome-wide significant loci. Independent SNPs, which had genome-wide significant associations with a Biomarker PC in either main effect, sex-stratified or sex interaction models.

**Table 3:** Mediation models. Results from structural equation models testing the hypothesis, that SNP effects on CSF biomarker PCs would also affect AD risk.

| SNP | Chr:Pos | Location | Locus | EA | EAF | R<sup>2</sup><sub>imp</sub> | Sex | PC1 | PC2 | PC3 | PC4 | PC5 | PC | Prop. | Direct | Total | Z<sub>AD</sub><br>(Wightman et al.) |
| --- | --- | --- | --- | --- | --- | --- | --- | --- | --- | --- | --- | --- | --- | --- | --- | --- | --- |
| *Tau pathology/Degeneration* |  |  |  |  |  |  |  |  |  |  |  |  |  |  |  |  |  |
| rs140169162 | 17:30886881 | Intron | MYO1D | G | 0.98 | 0.69 | m | 0.02 | 0.10 | -0.03 | 0.01 | 0.00 | 0.11 | 0.29 | 0.26 | 0.36 | -1.94 |
|  |  |  |  |  |  |  | f | 0.52* | -0.12 | -0.04 | -0.01 | 0.00 | 0.35* |  | -1.04* | -0.69 |  |
| *Aβ Pathology* |  |  |  |  |  |  |  |  |  |  |  |  |  |  |  |  |  |
| rs1638675 | 10:119178376 | Intergenic | - | A | 0.57 | 0.89 | m | 0.03 | 0.10* | -0.03 | 0.00 | 0.00 | 0.10* |  | -0.06 | 0.04 | -0.18 |
|  |  |  |  |  |  |  | f | 0.02 | -0.04* | 0.00 | 0.00 | 0.00 | -0.02 |  | 0.07 | 0.04 |  |
| *Tau pathology/Degeneration & Aβ Pathology* |  |  |  |  |  |  |  |  |  |  |  |  |  |  |  |  |  |
| rs12972156 | 19:45387459 | Intron | APOE | C | 0.74 | 0.98 | all | -0.07* | -0.12* | 0.02 | 0.00 | 0.00 | -0.17* | 0.65 | -0.09 | -0.27* | -Inf* |
| rs157582 | 19:45396219 | Intron | APOE | C | 0.65 | 1.00 | all | -0.08* | -0.11* | 0.01 | 0.00 | 0.00 | -0.17* | 0.79 | -0.05 | -0.22* | -Inf* |
| rs10119 | 19:45406673 | 3'-UTR | APOE | G | 0.60 | 0.95 | all | -0.06* | -0.10* | 0.01 | 0.00 | 0.00 | -0.15* | 0.67 | -0.07 | -0.23* | -Inf* |
| rs429358 | 19:45411941 | Exon | APOE | T | 0.70 | 0.99 | all | -0.09* | -0.13* | 0.01 | 0.00 | 0.00 | -0.21* | 0.54 | -0.18* | -0.39* | -Inf* |
| rs75627662 | 19:45413576 | Intergenic | APOE | C | 0.71 | 0.98 | all | -0.06* | -0.10* | 0.02 | 0.00 | 0.00 | -0.14* | 0.59 | -0.10 | -0.24* | -Inf* |
| rs157595 | 19:45425460 | Intergenic | APOE | A | 0.33 | 0.96 | all | -0.06* | -0.09* | 0.02 | 0.00 | 0.00 | -0.13* | 0.54 | -0.11 | -0.23* | -29.68* |
| rs111789331 | 19:45427125 | Intergenic | APOE | T | 0.73 | 0.95 | all | -0.08* | -0.12* | 0.02 | 0.00 | 0.01 | -0.19* | 0.57 | -0.14 | -0.33* | -Inf* |
| *Injury/Inflammation* |  |  |  |  |  |  |  |  |  |  |  |  |  |  |  |  |  |
| rs7551263 | 1:203150756 | Intron | CHI3L1 | T | 0.84 | 0.99 | all | 0.04 | -0.01 | 0.05* | -0.02 | 0.00 | 0.06 |  | -0.11 | -0.05 | 1.21 |
| rs114211800 | 5:158973225 | Intron | LOC105377684 | G | 0.98 | 0.91 | m | 0.11 | -0.08 | -0.31* | 0.00 | 0.00 | -0.29* |  | 0.27 | -0.02 | 0.45 |
|  |  |  |  |  |  |  | f | -0.09 | -0.05 | 0.03 | 0.01 | 0.00 | -0.09 |  | 0.58 | 0.49 |  |
| rs2302634 | 7:12270770 | Intron | TMEM106B | T | 0.58 | 1.00 | all | -0.02 | -0.01 | 0.07* | 0.00 | 0.00 | 0.04 |  | -0.08 | -0.04 | 5.38* |
| rs56194026 | 15:74831534 | Intergenic | ARID3B | C | 0.92 | 0.90 | m | -0.03 | 0.01 | -0.13* | 0.01 | 0.00 | -0.15* |  | 0.09 | -0.06 | -0.03 |
|  |  |  |  |  |  |  | f | -0.01 | -0.03 | 0.10* | 0.00 | 0.00 | 0.06 |  | -0.18 | -0.12 | -0.01 |
| *No evidence for mediation* |  |  |  |  |  |  |  |  |  |  |  |  |  |  |  |  |  |
| rs10399931 | 1:203156080 | Intergenic | CHI3L1 | T | 0.24 | 0.98 | all | 0.04 | -0.03 | 0.02 | -0.03 | 0.00 | -0.01 |  | 0.10 | 0.10 | 0.58 |
| rs12670437 | 7:76390507 | Intergenic | - | G | 0.98 | 0.87 | all | -0.10 | 0.05 | -0.01 | -0.01 | 0.00 | -0.07 | 0.54 | -0.06 | -0.14 | 0.22 |
| rs145791381 | 9:24796757 | Intergenic | - | T | 0.98 | 0.93 | all | 0.00 | -0.01 | 0.10 | -0.03 | 0.00 | 0.06 |  | -0.08 | -0.02 | 2.20* |
| rs16974493 | 13:110307768 | Intergenic | - | T | 0.97 | 0.98 | all | -0.01 | 0.01 | 0.01 | 0.02 | 0.00 | 0.03 |  | -0.07 | -0.03 | 0.34 |
| rs150326618 | 14:52641990 | Intergenic | - | G | 0.98 | 0.80 | all | -0.08 | -0.02 | -0.07 | 0.03 | 0.00 | -0.15 | 0.19 | -0.62 | -0.77 | -0.22 |
| rs275844 | 19:48884949 | Intergenic | *GRIN2D* | G | 0.88 | 0.93 | m | -0.07 | 0.02 | -0.01 | 0.00 | 0.00 | -0.06 |  | 0.18 | 0.12 | -2.13* |
|  |  |  |  |  |  |  | f | -0.02 | 0.05 | 0.02 | -0.01 | 0.00 | 0.05 |  | -0.18 | -0.13 |  |
| rs8111684 | 19:48947326 | 3'-UTR | *GRIN2D* | G | 0.26 | 1.00 | all | 0.02 | 0.00 | -0.02 | 0.00 | 0.00 | 0.00 | 0.04 | -0.01 | -0.01 | 0.72 |
| rs3107911 | 19:48964305 | Intron | *GRIN2D* | C | 0.15 | 0.96 | all | 0.02 | 0.00 | -0.01 | 0.00 | 0.00 | 0.00 | 0.03 | -0.05 | -0.05 | 1.31 |

**Table 4:** Gene-based tests. Results from gene-based tests with MAGMA representing the joint effects of common SNPs withing the named gene.

| Outcome | Gene | Chr | Start | End | N<sub>snp</sub>s | Z | p |
| --- | --- | --- | --- | --- | --- | --- | --- |
| *Genome-wide significant genes* |  |  |  |  |  |  |  |
| *Aβ Pathology* | *APOE* | 19 | 45409011 | 45412650 | 6 | 7.91 | 1.3E-15 |
| Injury/Inflammation | *TMEM106B* | 7 | 12250867 | 12282993 | 182 | 5.75 | 4.6E-09 |
| Non-AD Inflammation | *CHI3L1* | 1 | 203148059 | 203155877 | 28 | 4.53 | 2.9E-06 |
| Non-AD Synaptic Functioning | *GRIN2D* | 19 | 48898132 | 48948188 | 110 | 4.63 | 1.8E-06 |
| *Comparison with rare-variant hits* |  |  |  |  |  |  |  |
| Injury/Inflammation | *IFFO1* | 12 | 6647541 | 6665239 | 46 | 0.50 | 0.307 |
| Injury/Inflammation | *DTNB* | 2 | 25600067 | 25896503 | 385 | -1.11 | 0.867 |
| Injury/Inflammation | *NLRC3* | - | - | - | - | - | - |
| Injury/Inflammation | *SLC22A10* | 11 | 62905339 | 63137190 | 564 | -1.79 | 0.963 |
| Non-AD Synaptic Functioning | *GABBR2* | 9 | 101050391 | 101471479 | 1445 | 1.41 | 0.079 |
| Non-AD Synaptic Functioning | *CASZ1* | 1 | 10696661 | 10856707 | 387 | 0.32 | 0.374 |

**Chr** Chromosome
**Start/End** SNPs between start and end were considered
**N<sub>snp</sub>s** Number of SNPs included in test
**Z** Z test statistic
**p** p-value

### Main Analyses

In the main analyses, none of the SNPs showed genome-wide significant association with the tau pathology/degeneration component (PC1). In contrast, seven independent SNPs in the *APOE* locus showed genome-wide significant associations with the Aβ pathology component (PC2). Specifically, the C-allele of the lead SNP rs429358 (which is also known as the ε4-allele) was associated with −0.50SD lower PC scores (SE=0.50, p=1.3*10^−29^).

Regarding the injury/inflammation component (PC3), transmembrane protein 106B (*TMEM106B)*, tagged by lead intronic SNP rs2302634, showed strong and genome-wide significant associations: the T allele at this variant predicted +0.26SD (SE=0.04, p=1.3*10^−9^) higher injury/inflammation scores.

In case of the non-AD inflammation component (PC4) two loci reached genome-wide significance: chitinase 3 like 1 (*CHI3L1)* on chromosome 1 and a new region on chromosome 9p21.3 with lead SNP rs145791381 (located in an intergenic region). *CHI3L1* encodes the YKL-40 protein and two independent SNPs in or near this gene showed genome-wide significant associations with the non-AD inflammation PC: the strongest effect was observed for intronic variant rs7551263 (T allele: β=-0.39SD, SE=0.05, p=5.7*10^−17^; Figure 4). The second signal in *CHI3L* was elicited by SNP rs10399931 located <160bp upstream of *CHI3L* (T allele: β=-0.33SD, SE=0.06, p=6.0*10^−9^). Both SNPs are independent (r^2^=0.04 in our study and D’=0.71 in European populationl^40^) and therefore probably represent two separate *cis* pQTL signals. Another SNP reaching genome-wide significance in the main effect analyses was the intergenic SNP rs145791381 on chr. 9p21.3. The T allele of this variant was associated with lower scores on the non-AD inflammation component (PC4, β=-1.01SD, SE=0.18, p=6.0*10^−9^).

Lastly, no single SNP showed genome-wide significant association with the non-AD synaptic functioning component (PC5). However, this PC showed evidence for genome-wide significant association in the gene-based GWAS analyses highlighting the glutamate receptor gene *GRIN2D*, based on aggregated test statistics across 110 SNPs located between 19:48898132 and 19:48948188 (Z=+4.63, p=1.8*10^−6^) (Table 4). The lead SNP in this region was rs8111684 in the 3’ UTR region of the GRIN2D gene (β=+0.19SD, SE=0.05, p=9.8*10^−5^). Interestingly, the genomic regions flanking the gene both p-ter and q-ter showed more significant SNP associations. As these variants are located outside of the gene, they were not considered in the gene-based tests (Figure 4), we therefore included two additional SNPs in this region for further characterization: rs275844 and rs3107911. The strongest single SNP association in the locus was elicited by rs275844, located in the intron of the gene *KCNJ14* (β=-0.30SD, SE=0.07, p=9.6*10^−7^) and ca. 16kb q-ter of *GRIN2D*.

### Sex-specific effects

Several additional SNPs showed genome-wide significance only in the male or female subsamples. For instance, rs114211800, an intronic variant of the non-coding RNA gene *LOC105377684* on chromosome 5q33.3 showed strong association with the injury/inflammation component (PC3) in males (β=-1.23SD, SE=0.22, p=1.7*10^−8^), but not in females (β=-0.11SD, SE=0.26, p=0.69). Similarly, the intergenic SNP rs12670437 (chromosome 7q11.23) was strongly associated with non-AD inflammation in male participants (β=-1.20SD, SE=0.22, p=2.4*10^−8^), but not in females (β=+0.20SD, SE=0.21, p=0.33). Vice versa, rs140169162, located in an intron of the *MYO1D* gene on chromosome 17q11.2, showed highly specific effects on the component capturing tau pathology/degeneration (PC1) in females (β=+1.61SD, SE=0.30, p=4.9*10^−8^), but not in males (β=-0.02SD, SE=0.26, p=0.94). Likewise, for the component tagging non-AD inflammation (PC4), rs16974493 (intergenic, chr. 13q33.3) and rs150326618 (intergenic, chr. 14q22.1) were only genome-wide significant in females (rs16974493: β=-0.97SD, SE=0.17, p=2.2*10^−8^; rs150326618: β=+1.39SD, SE=0.25, p=2.6*10^−8^), but not in males (rs16974493: β=+0.18SD, SE=0.22, p=0.42; rs150326618: β=+0.09SD, SE=0.24, p=0.72).

In addition to these sex-specific results, the sex interaction models revealed two SNPs eliciting significant evidence for sex interaction reflecting their opposite effects in males vs. females: rs1638675 (intergenic, chr. 10q25.3) and rs56194026 (approx. 2kb upstream of *ARID3B* on chr. 15q24.1). For SNP rs1638675, the A allele showed a negative association with Aβ pathology in males (β=-0.33SD, SE=0.07, p=1.0*10^−6^), but positive in females (β=+0.18SD, SE=0.06, p=4.2*10^−3^). In case of rs56194026, the association was sex-dependent for the injury/inflammation component (PC3) where the C allele showed a negative effect in males (β=-0.50SD, SE=0.12, p=1.6*10^−5^), but a positive effect in females (β=+0.41SD, SE=0.12, p=5.3*10^−4^).

Finally, we highlight a suggestive sex difference for rs2302634 in *TMEM106B*. The effect of this SNP on the component capturing injury/inflammation (PC3) was approximately twice as large in males compared to females (β_male_=+0.34SD vs β_female_=+0.18SD), although this difference did not attain statistical significance (p=0.06).

### Mediation analyses

Our main GWAS main analyses identified several loci showing highly significant association with the biomarker PCs defined for this study. As three of the five biomarker PCs are independently associated with diagnostic status, this raises the questions as to whether the SNP effects on PC levels also significantly impact AD development. Overall, we observed two distinct mediation patterns: 1.) SNPs that affect AD either via alteration in both the Aβ pathology and tau pathology/degeneration components (*APOE*) or 2.) SNPs that affect AD via the injury/inflammation PC only (*TMEM106B* and *CHI3L1*).

In the case of *APOE*, the rs429358 ε4 allele was associated with a 0.39SD higher latent AD score (SE=0.06, p=4.6*10^−12^). The mediation model suggests, that this adverse effect can be partitioned into three pathways: 1.). One third is attributable to mediation via the Aβ pathology component (PC2), 2.) 23% of the SNP effects were due to mediation via the tau pathology/neurodegeneration component (PC1), while 3.) the remaining 46% were due to pathways not represented by any of the measured biomarker PCs (Table 3).

The mediation analyses also suggested, that increases in injury/inflammation scores (PC3) due to the T allele in SNP rs2302634 (*TMEM106B)* resulted in a significant increase of latent AD (β=+0.07SD, SE=0.02, p=1.2*10^−5^). Furthermore, we did not find evidence, that *TMEM106B* affects AD by any other pathway, either measured or unmeasured. The positive mediation effect is consistent with the strong positive total AD risk effect for rs2302634 (+Z=5.38, p=7.3*10^−8^), and overall genome-wide significant association with AD risk recently described by two GWAS.^41,42^ See Figure 4 for a regional plot visualizing associations between *TMEM106B* with the biomarker PC and previously reported associations with AD.

The SNP rs7551263 in the intron of *CHI3L1* was primarily associated with the component capturing non-AD inflammation (PC4). As this PC did not correlate with latent AD, we also found no evidence for mediation of AD risk via this pathway. However, rs7551263 was also nominally associated with the injury/inflammation component (PC3) (T allele: β=+0.21SD, SE=0.06, p=2.3*10^−4^) and showed evidence for mediation through this pathway (β=+0.05SD, SE=0.02, p=4.0*10^−4^). Interestingly, the T allele was negatively associated with non-AD inflammation (PC4), so our results suggest that while the T allele decreases levels of non-AD inflammation biomarker profiles this has no measurable protective effect on latent AD. At the same time, this allele significantly increases injury/inflammation profiles, which results in a significantly higher AD risk.

### Comparison to rare variant results

In contrast to our previous work based on WES-derived rare variants in a subset of the EMIF-MBD dataset analyzed here,^3^ we found no evidence for an association between the analogous CSF biomarker components and common variants in the genes previously highlighted (i.e., *IFFO1*, *DTNB*, *NLRC3, SLC22A10*, *GABBR2* and *CASZ*). Similarly, combining common SNP effects in gene-based tests did not reveal any significant associations at these loci either (Table 4). Together, these results suggest that common variants (MAF ≥0.01) do not appreciably contribute to the rare variant association signals identified earlier by our group.

## Discussion

In this work, we comprehensively explored the influence of common variants on multivariate combinations of AD CSF biomarkers representing different disease processes. In addition to confirming several previously reported GWAS loci, we identified two new regions (*GRIN2D* and rs145791381) showing strong association with synaptic functioning and inflammation in AD. Furthermore, our results provide evidence for the presence of numerous loci with sex-specific effects.

Arguably the most interesting finding of our main GWAS analyses is the discovery of genome-wide significant gene-based association with variants in *GRIN2D* on chromosome 19q13.33 and the non-AD synaptic functioning component (PC5). Interestingly, SNPs in the chromosomal regions immediately flanking *GRIN2D* showed an even stronger association with non-AD synaptic functioning than variants within *GRIN2D* itself, possibly suggesting that gene expression rather than gene (dys)functioning may be the lead mechanism underlying this association. *GRIN2D* encodes the GluN2D subunit of the glutamate receptor NMDAR, which plays an important role in learning and memory.^43^ While mutations in *GRIN2D* have been reported to cause epileptic encephalopathy,^43^ this gene’s role in AD and other traits is less clear. While some recent data suggest that *GRIN2D* mRNA expression is lower in the temporal cortex of AD cases according to the AMP-AD project,^44^ there are no strong GWAS-based association signals reported in this region of chromosome 19 and relevant cognitive traits in the GWAS catalog (https://www.ebi.ac.uk/gwas/regions/chr19:48361628-48476971), except for an association with self-reported mathematical ability. Look-up of our two lead variants in the *GRIN2D* region (i.e. rs275844 and rs3107911) in summary statistics of two recent AD GWAS suggest a weak association for rs275844 in only one of the GWAS (p=0.03)^41^. Our mediation analyses did not identify significant effects on latent AD, either, suggesting no or weak association of the locus with AD. In conclusion, we found strong and novel evidence of a role of *GRIN2D* in non-AD related synaptic functioning in the CSF biomarker component mainly driven by Ng.

The second most interesting signal of our main GWAS analyses was the association between SNP rs145791381 and the non-AD inflammation component (PC4), which represents YKL-40 levels not associated with other AD biomarkers or AD status. The SNP is located in a genepoor region, is not very common (MAF=2%), and not mentioned as a GWAS signal in previous literature (e.g. the GWAS catalog). It is therefore difficult to speculate about potential mechanisms or to judge the plausibility of this finding. While further research is needed to confirm and interpret this finding, it is noteworthy to mention that in sex stratified analyses the SNP showed highly consistent effect sizes and genome-wide significance in both sexes, suggesting a robust finding.

Besides these novel associations, the current study also provides further insights into how known AD loci affect disease risk by quantifying the biomarker mediated risk effects. Overall, we observed two CSF biomarker profiles associated with AD, which are determined by two different gene sets. The first profile (PC1 and PC2) is characterized by decreased amyloid and increased tau, as well as increased Ng and YKL-40 levels, but not NfL. This component was most strongly associated with SNPs in the *APOE* region, in particular the well-known AD risk variant ε4. Our data suggest that the association with this variant may increase AD risk by being a catalyst for amyloid deposition or as inhibitor of amyloid clearance, represented here by the Aβ pathology PC. The resulting amyloid aggregation is thought to cascade into several neurodegenerative processes, involving formation of tau tangles, loss of synaptic functioning and inflammation.^45^ Astonishingly, our results suggest that combinations of Aβ, tau, Ng and YKL-40 assessments are able to capture most of these neurodegenerative processes triggered by the *APOE* locus, as they mediated 54-79% of the APOE SNP effects. However, it is important to note that the four CSF biomarkers captured by PC1 and PC2 are not sufficient to explain all genetic risk effects on AD. The second most relevant CSF biomarker pattern in that regard was the injury/inflammation component (PC3) represented by increased NfL and YKL-40 levels. PC3 levels are statistically independent of the changes in amyloid and tau levels (captured by PC1 and PC2), typically observed in AD, but associated with AD diagnostic status to a similar strong degree. Given prior knowledge of the non-specificity of NfL with respect to AD pathogenesis,^6^ we interpret this pattern to represent an independent non-AD-specific neurodegenerative pathway for dementias in general. Genetically, our results suggest that this pathway is not determined by variants in the *APOE* locus, but instead by variants in *TMEM106B* and potentially *CHI3L1. TMEM106B* affects neuronal loss^46^ and has been convincingly associated with risk for at least two forms of dementias, i.e. fronto-temporal dementia^46^ and AD^13,41,47^. *CHI3L1* encodes the YKL-40 protein and we and others have previously demonstrated that common and possibly rare variants at this locus represent *cis* pQTLs of CSF YKL-40.^3,13^ Our results suggest that *CHI3L1* variants may be associated with increased neuronal injury and inflammation leading to a heightened AD risk, however, the association was not as strong, as with inflammation variance independent of AD.

Additional insights resulted from the sex-stratified analyses which revealed several SNPs either showing associations in only one sex stratum, or opposite effects in males and females. As examples we highlight two such SNPs: First, rs140169162 is located in an intron of *MYO1D* and showed strong association with tau pathology/neurodegeneration (PC1) with evidence for a mediation effect on latent AD, but only in the female subsample. Interestingly, SNPs in *MYO1D* has been found to have a female-specific effect on hernias, as well.^48^ Despite its apparent sex-specificity previous work has nominated *MYOD1* as a potential drug target for AD according to predictive network analysis.^49,50^ The encoded protein, myogenic differentiation 1, is involved in myelin sheath formation^51^ and both common^52^ and rare^53^ variants have been associated with autism, supporting *MYO1D’s* role in neural development and functioning.

Second, rs56194026 is located near *ARID3B* and was associated with injury/inflammation (PC3), with strong but opposite effects in males vs. females. The gene encodes AT-rich interaction domain 3B, a DNA-binding protein from the ARID family of proteins which are involved in embryonic patterning, cell lineage gene regulation, cell cycle control, transcriptional regulation and possibly in chromatin structure modification.^54^ Samyesudhas et al.^55^ recently suggested a relevant role of this protein in AD development, as *ARID3B* is expressed in response to the amyloid precursor protein intracellular domain and neuronal injury. However, it remains unclear, why SNPs near this gene would have opposite effects in males and females. Possibly, this is related to the higher mean NfL levels in males, or the genes’ proposed function as regulator of sex-biased expression^56^

A major strength of our study is the application of multivariate analyses based on five CSF biomarker profiles and the estimation of mediation effects. Studying component patterns of different biomarker combinations allows to shed new light and provide new insights on how common genetic variants affect biomarkers and AD risk beyond their effects on the levels of single biomarkers. The fact that this distinction can be quite important is evidenced by our results with YKL-40 and Ng which show different association patterns, depending on whether or not they co-vary with the levels of other biomarkers. The inclusion of the X-chromosome and examination of sex-differences are additional strengths of our study. While no SNPs on the X-chromosome attained genome-wide significance, we identified several SNPs showing sex-specific effects. This highlights the importance of modeling sex interactions, especially for biomarkers with pronounced differences in mean levels.

In addition to these strengths, we note the following potential limitations. First, while our sample size is generally large for a CSF biomarker study, it is small compared to GWAS of other complex traits, including recent GWAS in the AD field.^41,42^ Second, the sample size limitation is aggravated in the sex-specific analyses that need to be interpreted with caution and require further replication. Third, we only studied individuals of European descent. It remains unclear whether and to which degree our results are relevant also in non-European ancestries. It is well known, for instance, that the *APOE* risk effects on AD are ancestry-dependent.^57,58^ Fourth, it is important to emphasize, that our mediation analyses are based on the assumption that the analyzed CSF biomarkers reflect pathological processes that *precede* and *cause* AD symptoms. An alternative – and often times equally plausible – interpretation is that the uncovered SNPs affect AD symptoms independently of biomarker levels and that the component associations observed here actually reflect a *consequence* of AD pathogenesis. While the specificity of the mediation results using certain PCs but not others generally supports the assumed causal directions, longitudinal studies, e.g., on MCI conversion, are needed to confirm the findings of this arm of our project.

In summary, in this first multivariate CSF biomarker GWAS we observed two novel loci showing strong and convincing association with non-AD specific biomarker patterns: *GRIN2D* with non-AD synaptic functioning and rs145791381 with non-AD inflammation. The results also suggest the presence of two distinct mediation pathways, by which common SNPs may affect AD risk. One pathway is related to amyloid and tau pathology and is mostly determined by *APOE* SNPs. The second pathway is related to increased neuronal injury and inflammation, captured by NfL and YKL-40. Genetically, this latter pathway is mostly driven by variants in *TMEM106B* and *CHI3L1*. Pathway-aware genetic studies with larger sample sizes and in more diverse ancestries are needed to further understand the complex etiology of AD and to translate genetic information to personalized medicine approaches.

## Data Availability

In accordance with EU law and participant privacy, clinical individual-level data from EMIF-AD is not available publicly, but can be obtained via EMIF-AD (https://emif-catalogue.eu;http://www.emif.eu/about/emif-ad). Registered users can download ADNI data from http://adni.loni.usc.edu/. Analysis code can be found at https://github.com/aneumann-science/common_variants_csf_biomarkers. Summary statistics for all GWAS will be made publicly available after publication.

https://emif-catalogue.eu

http://www.emif.eu/about/emif-ad

http://adni.loni.usc.edu/

## Acknowledgments

**EMIF**

We thank the study participants and their families, as well as all people, who contributed data and sample collection and/or logistics across the different study centres.

Funding: The present study was conducted as part of the EMIF-AD project, which has received support from the Innovative Medicines Initiative Joint Undertaking under EMIF grant agreement no. 115372, EPAD grant no. 115736, and from the European Union’s Horizon 2020 research and innovation programme under grant agreement No. 666992. Resources of which are composed of financial contribution from the European Union’s Seventh Framework Program (FP7/2007-2013) and EFPIA companies’ in-kind contribution. Research at VIB-UAntwerp was in part supported by the Research Foundation Flanders (FWO), and the University of Antwerp Research Fund, Belgium. The DESCRIPA study was funded by the European Commission within the fifth framework program (QLRT-2001-2455). The EDAR study was funded by the European Commission within the fifth framework program (contract no. 37670). The San Sebastian GAP study is partially funded by the Department of Health of the Basque Government (allocation 17.0.1.08.12.0000.2.454.01.41142.001.H). The Leuven cohort was funded by the Stichting voor Alzheimer Onderzoek (grant numbers #11020, #13007 and #15005). The Lausanne cohort was supported by grants from the Swiss National Research Foundation (SNF 320030_141179), Synapsis Foundation-Alzheimer Research Switzerland (grant number 2017-PI01). HZ is a Wallenberg Scholar supported by grants from the Swedish Research Council (#2018-02532), the European Research Council (#681712 and #101053962), Swedish State Support for Clinical Research (#ALFGBG-71320), the Alzheimer Drug Discovery Foundation (ADDF), USA (#201809-2016862), the AD Strategic Fund and the Alzheimer’s Association (#ADSF-21-831376-C, #ADSF-21-831381-C and #ADSF-21-831377-C), the Bluefield Project, the Olav Thon Foundation, the Erling-Persson Family Foundation, Stiftelsen för Gamla Tjänarinnor, Hjärnfonden, Sweden (#FO2022-0270), the European Union’s Horizon 2020 research and innovation programme under the Marie Skłodowska-Curie grant agreement No 860197 (MIRIADE), the European Union Joint Programme – Neurodegenerative Disease Research (JPND2021-00694), and the UK Dementia Research Institute at UCL (UKDRI-1003). FB is supported by the NIHR biomedical research centre at UCLH. Further support came from the Lifebrain EU Horizon 2020 project (#732592, to CML and LB), the Deutsche Forschungsgemeinschaft (#LI2654/2-1 and LI 2654/4-1 to CML and #BE2287/6-1 to LB), and the Cure Alzheimer’s Fund (to CML and LB). Lastly, we acknowledge support from the high-performance environment (“OmicsCluster”) at University of Lübeck where most of the GWAS-related analyses were performed.

**ADNI**

Data used in preparation of this article were obtained from the Alzheimer’s Disease Neuroimaging Initiative (ADNI) database (adni.loni.usc.edu). As such, the investigators within the ADNI contributed to the design and implementation of ADNI and/or provided data but did not participate in analysis or writing of this report. A complete listing of ADNI investigators can be found at: http://adni.loni.usc.edu/wp-content/uploads/how_to_apply/ADNI_Acknowledgement_List.pdf. Data collection and sharing for this project was funded by the Alzheimer’s Disease Neuroimaging Initiative (ADNI) (National Institutes of Health Grant U01 AG024904) and DOD ADNI (Department of Defense award number W81XWH-12-2-0012). ADNI is funded by the National Institute on Aging, the National Institute of Biomedical Imaging and Bioengineering, and through generous contributions from the following: AbbVie, Alzheimer’s Association; Alzheimer’s Drug Discovery Foundation; Araclon Biotech; BioClinica, Inc.; Biogen; Bristol-Myers Squibb Company; CereSpir, Inc.;Cogstate;Eisai Inc.; Elan Pharmaceuticals, Inc.; Eli Lilly and Company; EuroImmun; F. Hoffmann-La Roche Ltd and its affiliated company Genentech, Inc.; Fujirebio; GE Healthcare; IXICO Ltd.; Janssen Alzheimer Immunotherapy Research & Development, LLC.; Johnson & Johnson Pharmaceutical Research & Development LLC.; Lumosity; Lundbeck; Merck & Co., Inc.; Meso Scale Diagnostics, LLC.;NeuroRx Research; Neurotrack Technologies;Novartis Pharmaceuticals Corporation; Pfizer Inc.; Piramal Imaging; Servier; Takeda Pharmaceutical Company; and Transition Therapeutics. The Canadian Institutes of Health Research is providing funds to support ADNI clinical sites in Canada. Private sector contributions are facilitated by the Foundation for the National Institutes of Health (www.fnih.org). The grantee organization is the Northern California Institute for Research and Education, and the study is coordinated by the Alzheimer’s Therapeutic Research Institute at the University of Southern California. ADNI data are disseminated by the Laboratory for Neuro Imaging at the University of Southern California.

