## Supplementary Methods, Figure S1-S3, Table S1-S3 for "Multivariate GWAS of Alzheimer’s disease CSF biomarker profiles implies GRIN2D in synaptic functioning"

*X-chromosomal genotyping, imputation, and quality control (QC)*

For genotypes on the X chromosome, QC for non-pseudoautosomal regions markers was performed separately for male and female subjects. SNPs were excluded with MAF below 1% in the female group, genotyping rates below 98%, deviations from HWE in control individuals ($p<1\times10^{-4}$), differential genotyping efficiency between women and men ($p< 1\times10^{-4}$), differential allele frequency between women and men ($p< 1\times10^{-4}$) or ambiguous allele combinations (A/T and C/G). Markers in the pseudoautosomal regions (PAR1, PAR2) were processed analogous to the autosomes. We then aligned the alleles of the remaining SNPs to the reference genome "GRCh37/hg19" before imputation. Imputations of untyped X-Chromosomal markers (including PAR) were subsequently performed on the Sanger Institute imputation server (https://imputation.sanger.ac.uk/) using the extended HRC reference panel available at that site.[1] After imputation, we used the same QC criteria as for the autosomal SNPs on markers in PAR. For remaining non-PAR markers SNPs were excluded with MAF below 1% in male and female separately, imputation quality ($R^2<0.30$) and deviations from HWE ($p<5\times10^{-4}$).

*Multiple testing adjustment*

To strike a balance between reliable inference and power, we present our findings as primary, secondary and tertiary results. The primary analyses in this study were the GWAS in the full dataset independent of sex. For SNP-based tests we applied the conventional genome-wide association threshold of $p<5*10^{-8}$ and for gene-based tests we used Bonferroni's method to adjust for 19,511 genes resulting in a threshold of $p<2.3*10^{-6}$, as recommended by FUMA. The sex-specific analyses present additional tests of related (and non-independent) hypothesis, and, thus should be regarded as secondary and more exploratory analyses.

Next, we aimed to better understand the potential consequences of identified loci on latent AD and selected them for further mediation analyses. Here, we assume that the genome-wide significant SNPs are truly associated with a biomarker PC and test the hypothesis, that they are also associated with latent AD. As each SNP can be associated with latent AD either directly or via any of the five biomarker PCs, we adjusted for 6 possible pathways, resulting in a Bonferroni-adjusted alpha of $0.05/6=p<0.0083$.

In case of comparisons to rare variants, we tested SNPs based on prior evidence of rare variants associations. We therefore only adjusted for number of SNPs within the selected loci. To account for non-independence between SNPs, we adjusted for the number of effective tests, taking into account the eigenvalue of correlation between SNPs. We estimated the number of effective tests using the Li & Ji method[2], as implemented in poolR[3]. In the current study, we assessed 564 SNPs within *IFFO1*, *DTNB*, *NLRC3* and *SLC22A10* for association with the injury/inflammation component (PC3). The estimated number of effective tests was 127, resulting in a Bonferroni-adjusted alpha of $0.05/127=3.9*10^{-4}$. In case of *GABBR2* and *CASZ,* which were tested for association with non-AD synaptic functioning (PC5), we analyzed 1837 common genetic variants within both genes, 312 of which were independent resulting in a Bonferroni adjusted alpha of $p<0.05/312=1.6*10^{-4}$.

*References*

1. McCarthy, S., Das, S., Kretzschmar, W., Delaneau, O., Wood, A.R., Teumer, A., Kang, H.M., Fuchsberger, C., Danecek, P., Sharp, K., et al. (2016). A reference panel of 64,976 haplotypes for genotype imputation. Nat. Genet. *48*, 1279–1283.

2. Li, J., and Ji, L. (2005). Adjusting multiple testing in multilocus analyses using the eigenvalues of a correlation matrix. Heredity (Edinb). *95*, 221–227.

3. Cinar, O., and Viechtbauer, W. meff: Number of Effective Tests.

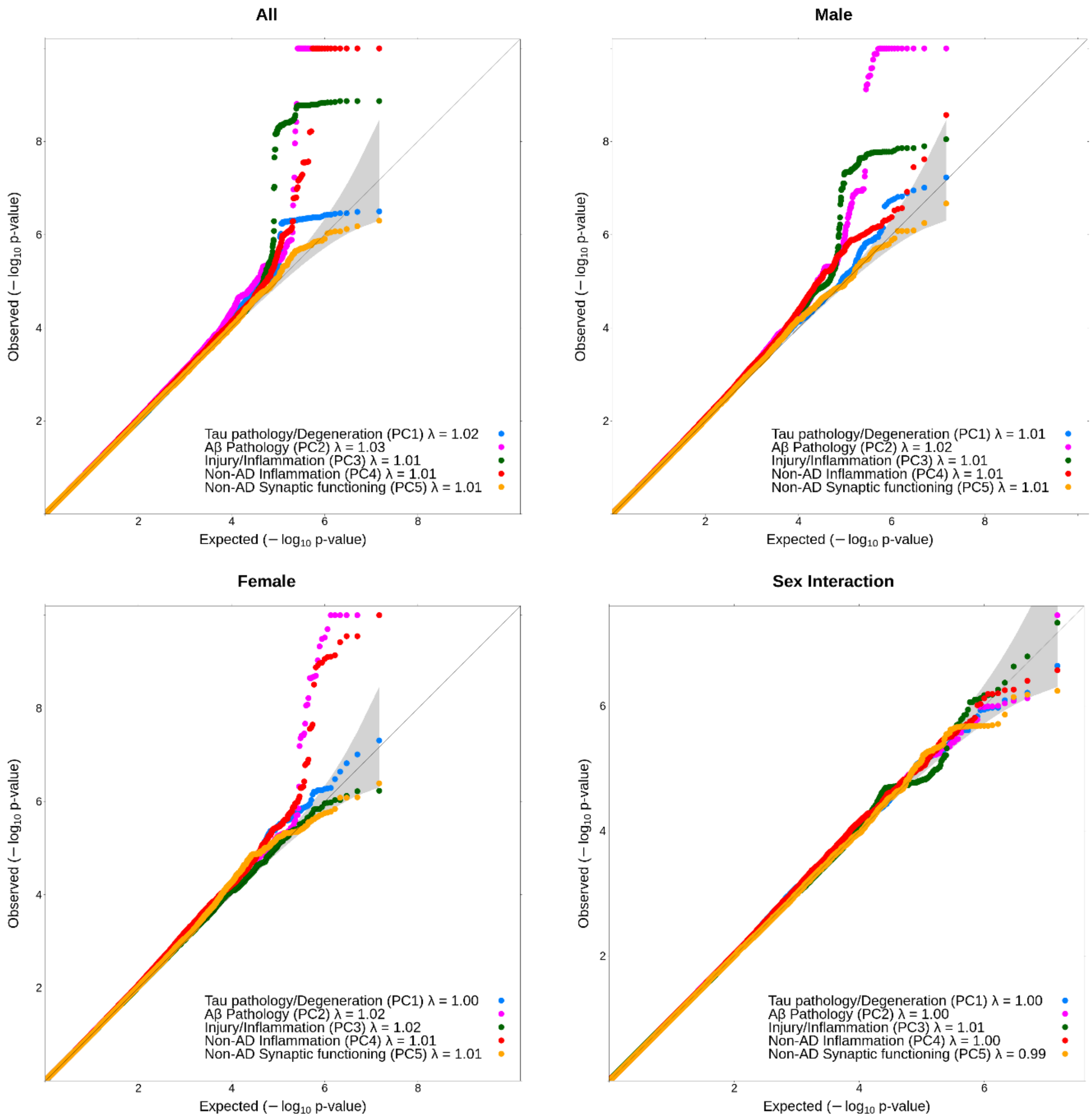


**Figure S1: QQ-plots of SNP test statistics.** Diagonal line indicates a p-value distribution expected by chance. Dots represent p-values per SNP for each of the outcomes (see legend for color coding). Each panel depicts the p-value distribution for different GWAS models, either main effects (all sexes), male or female-stratified, or the p-values of a sex interaction term.

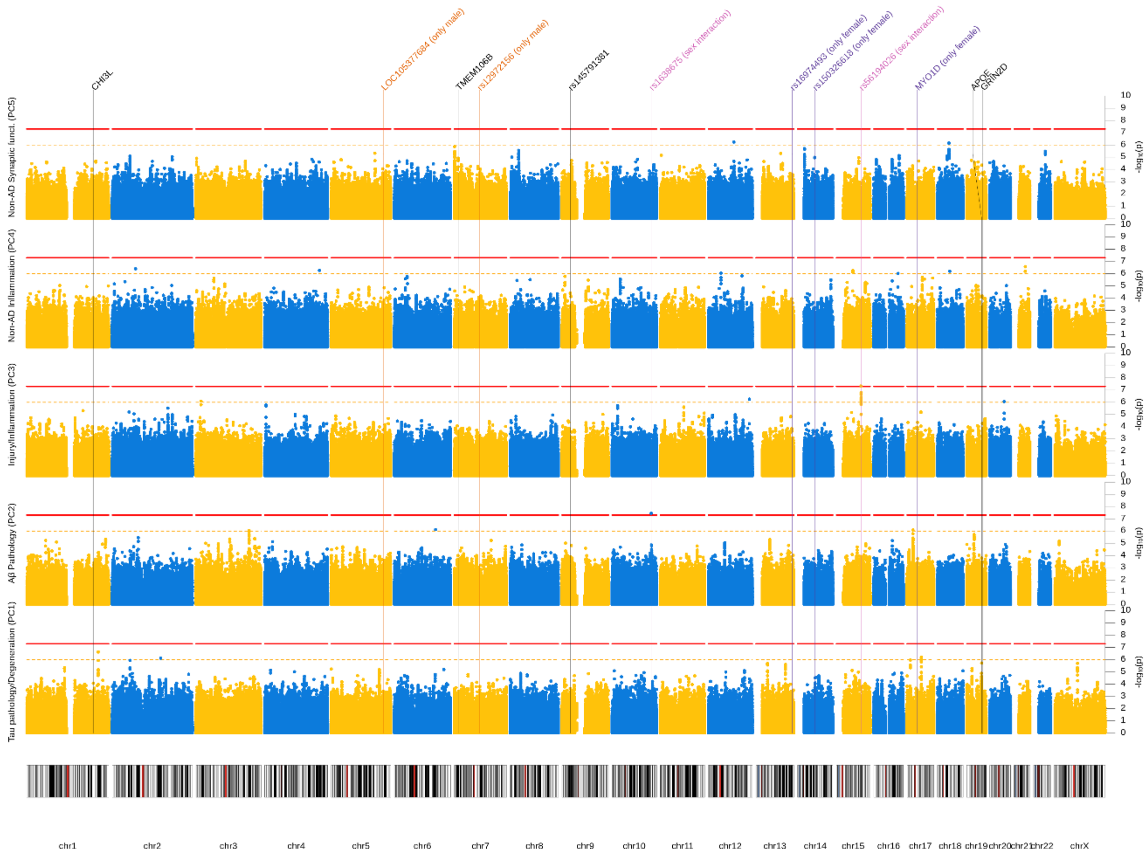


**Figure S2: Manhattan plot (sex interaction).** Sex-interaction results from GWAS of five CSF biomarker PC. Each row represents a different PC as outcome. X-axis represents each SNP and the y-axis the p-value of the sex-interaction term on a -$\log_{10}$ scale. All analyses were adjusted for genetic ancestry and SNP array. Red line indicates genome-wide significance threshold ($p=5*10^{-8}$). Yellow line indicates suggestive threshold ($p=1*10^{-6}$). Vertical lines point towards genome-wide significant loci based on any model. P-values below $1*10^{-10}$ were winsorized to $1*10^{-10}$

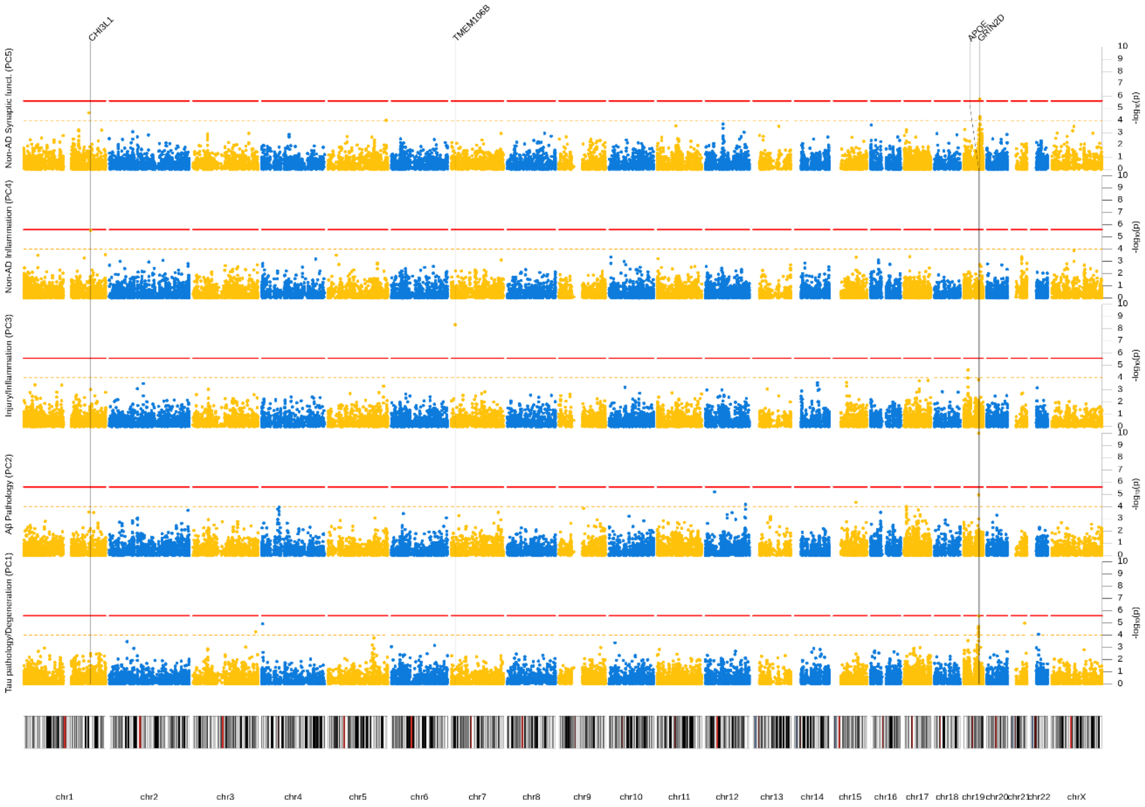


**Figure S3: Manhattan plot (gene-based tests).** Results from GWAS of five CSF biomarker PC across both sexes. Each row represents a different PC as outcome. X-axis represents each SNP and the y-axis the p-value of the gene-based association derived with MAGMA on a $-\log_{10}$ scale. All analyses were adjusted for sex, genetic ancestry, and SNP array. Red line indicates genome-wide significance threshold ($p=2.3*10^{-6}$). Yellow line indicates suggestive threshold ($p=1*10^{-4}$). Vertical lines point towards genome-wide significant loci based on any model. P-values below $1*10^{-10}$ were winsorized to $1*10^{-10}$.

### Table S1: Participant Characteristics CSF Biomarker Sample

| | EMIF (n=769) | | | | | | ADNI (n=389) | | | | | |
|---|---|---|---|---|---|---|---|---|---|---|---|---|
| | Male (n=370) | | | Female (n=399) | | | Male (n=231) | | | Female (n=158) | | |
| Characteristic | n | Mean/% | SD | n | Mean/% | SD | n | Mean/% | SD | n | Mean/% | SD |
| Age | 370 | 68.84 | 8.54 | 399 | 69.76 | 8.16 | 231 | 75.37 | 7.02 | 158 | 73.89 | 7.18 |
| Education (in years) | 370 | 11.77 | 3.73 | 399 | 10.24 | 3.61 | 231 | 16.11 | 3.07 | 158 | 14.89 | 2.81 |
| *Diagnosis* | | | | | | | | | | | | |
| Cognitively normal | 31 | 8.4% | - | 44 | 11.0% | - | 55 | 23.8% | - | 55 | 34.8% | - |
| Subjective cognitive impairment | 37 | 10.0% | - | 27 | 6.8% | - | 0 | 0 | - | 0 | 0% | - |
| Mild cognitive impairment | 215 | 58.1% | - | 234 | 58.6% | - | 123 | 53.2% | - | 62 | 39.2% | - |
| Alzheimer's disease | 87 | 23.5% | - | 93 | 23.3% | - | 53 | 22.9% | - | 41 | 26.0% | - |
| Mini-mental state examination | 369 | 25.94 | 3.61 | 158 | 25.16 | 4.21 | 231 | 26.68 | 2.54 | 158 | 26.79 | 2.68 |
| *CSF biomarkers* | | | | | | | | | | | | |
| β-Amyloid$_{42}$ (in pg/ml) | 369 | 324.71 | 198.80 | 397 | 1140.84 | 2242.28 | 231 | 874.81 | 440.57 | 158 | 951.68 | 455.24 |
| Tau (in pg/ml) | 368 | 359.07 | 305.75 | 397 | 405.16 | 364.22 | 231 | 294.22 | 122.39 | 158 | 296.61 | 148.87 |
| Phosphorylated Tau (in pg/ml) | 369 | 57.74 | 32.71 | 399 | 61.93 | 35.13 | 231 | 28.98 | 14.15 | 158 | 28.74 | 15.87 |
| Neurofilament light (in pg/ml) | 366 | 1306.58 | 1920.92 | 395 | 1140.84 | 2242.28 | 231 | 1582.10 | 1160.31 | 158 | 1277.99 | 527.32 |
| YKL-40 (in pg/ml or z-score*) | 370 | 171469.09 | 64960.25 | 398 | 173950.23 | 65679.59 | 52 | 0.17 | 0.97 | 34 | -0.09 | 0.89 |
| Neurogranin (in pg/ml) | 333 | 136.74 | 196.51 | 158 | 147.84 | 153.24 | 231 | 440.00 | 356.00 | 158 | 436.51 | 307.58 |

**Table 1: Participant characteristics.** Demographic information and descriptive statistics for participants with information on at least four biomarkers.

**n** Sample size

**SD** Standard Deviation

***** For ADNI, average values of two peptide sequences, with two ion frequencies each, were averaged after z-score standardization

### Table S2: Participant Characteristics GWAS Sample

| | EMIF (n=672) | | | | | | ADNI (n=301) | | | | | |
|---|---|---|---|---|---|---|---|---|---|---|---|---|
| | Male (n=322) | | | Female (n=350) | | | Male (n=183) | | | Female (n=118) | | |
| Characteristic | n | Mean/% | SD | n | Mean/% | SD | n | Mean/% | SD | n | Mean/% | SD |
| Age | 322 | 68.85 | 8.52 | 350 | 70.05 | 8.15 | 183 | 75.38 | 6.79 | 118 | 73.91 | 7.33 |
| Education (in years) | 322 | 11.90 | 3.66 | 350 | 10.34 | 3.61 | 183 | 15.71 | 3.01 | 118 | 15.97 | 2.99 |
| *Diagnosis* | | | | | | | | | | | | |
| Cognitively normal | 27 | 9.9% | - | 23 | 10.6% | - | 43 | 23.5% | - | 43 | 36.4% | - |
| Subjective cognitive impairment | 32 | 8.4% | - | 37 | 6.6% | - | 0 | 0% | - | 0 | 0% | - |
| Mild cognitive impairment | 190 | 59.0% | - | 210 | 60.0% | - | 96 | 41.6% | - | 50 | 42.4% | - |
| Alzheimer’s disease | 73 | 22.7% | - | 80 | 22.9% | - | 44 | 19.0% | - | 25 | 21.2% | - |
| Mini-mental state examination | 321 | 25.87 | 3.65 | 349 | 25.17 | 4.23 | 183 | 26.72 | 2.47 | 118 | 27.14 | 2.63 |
| *CSF biomarkers* | | | | | | | | | | | | |
| β-Amyloid$_{42}$ (in pg/ml) | 322 | 316.29 | 196.75 | 348 | 298.63 | 177.71 | 183 | 819.00 | 441.05 | 118 | 893.79 | 451.59 |
| Tau (in pg/ml) | 320 | 370.51 | 309.19 | 348 | 415.34 | 376.82 | 183 | 304.36 | 118.37 | 118 | 314.37 | 148.90 |
| Phosphorylated tau (in pg/ml) | 321 | 59.05 | 32.90 | 250 | 62.59 | 35.87 | 183 | 29.81 | 13.40 | 118 | 30.81 | 16.19 |
| Neurofilament light (in pg/ml) | 318 | 1247.64 | 1736.51 | 346 | 1173.67 | 2386.09 | 183 | 1554.11 | 1247.24 | 118 | 1285.79 | 811.96 |
| YKL-40 (in pg/ml or z-score*) | 322 | 169938.49 | 64617.47 | 349 | 174465.75 | 66691.41 | 41 | 0.13 | 1.01 | 32 | -0.18 | 1.01 |
| Neurogranin (in pg/ml) | 289 | 136.72 | 196.75 | 329 | 149.32 | 157.89 | 183 | 469.03 | 297.35 | 118 | 504.47 | 306.39 |

**Table 2: Participant characteristics.** Demographic information and descriptive statistics for main GWAS analysis sample.

**n** Sample size

**SD** Standard Deviation

***** For ADNI, average values of two peptide sequences, with two ion frequencies each, were averaged after z-score standardization

#### Table S3: Sex differences in CSF biomarker PCs and associations with Alzheimer’s disease

| Outcome | Predictor | β | SE | p |
|---|---|---|---|---|
| *Sex differences in mean levels* | | | | |
| Tau pathology/Degeneration | Female | 0.14 | 0.06 | 1.74E-02 |
| Injury/Inflammation | Female | -0.01 | 0.06 | 8.04E-01 |
| Injury/Inflammation | Female | -0.40 | 0.05 | 2.66E-13 |
| Non-AD Inflammation | Female | 0.02 | 0.06 | 7.88E-01 |
| Non-AD Synaptic Functioning | Female | 0.21 | 0.06 | 6.37E-04 |
| *Latent AD regressed on PCs and Sex* | | | | |
| Latent AD | Female | 0.05 | 0.07 | 4.75E-01 |
| Latent AD | Tau pathology/Degeneration | 0.41 | 0.05 | 6.83E-17 |
| Latent AD | Tau pathology/Degeneration*Female | 0.01 | 0.07 | 8.56E-01 |
| Latent AD | Aβ Pathology | -0.34 | 0.05 | 1.09E-12 |
| Latent AD | Aβ Pathology*Female | -0.11 | 0.07 | 1.33E-01 |
| Latent AD | Injury/Inflammation | 0.40 | 0.06 | 2.75E-12 |
| Latent AD | Injury/Inflammation*Female | 0.05 | 0.07 | 5.12E-01 |

| | | | | |
|---|---|---|---|---|
| Latent AD | Non-AD Inflammation | 0.12 | 0.05 | 1.94E-02 |
| Latent AD | Non-AD Inflammation*Female | 0.04 | 0.07 | 5.70E-01 |
| Latent AD | Non-AD Synaptic Functioning | 0.04 | 0.05 | 4.17E-01 |
| Latent AD | Non-AD Synaptic Functioning*Female | -0.12 | 0.07 | 1.02E-01 |

**Table 4: Sex differences in CSF biomarker PCs and associations with Alzheimer's disease.**

**Rows 2-7** represent linear models, in which CSF Biomarker PCs are regressed on sex (Male=0, Female=1), adjusted for age, study and genetic ancestry components. Each outcome is fitted in a separate model. **β** represents the covariate adjusted difference in mean levels between females and males.

**Rows 9-19** represents a structural equation model in which a latent AD factor is regressed on sex, biomarker PCs, the interaction between sex and Biomarker PCs, as well as age, study and genetic ancestry components. **β** represents either the effect of being female on latent AD in SD assuming mean PC levels (Female), the effect of biomarker PCs on latent AD in males (biomarker PC), and the additional effect of the biomarker for females (biomarker PC*Female).

**SE** Standard Error

**p** p-value
